# Barriers to Access to Care Evaluation Scale - Proxy Report (BACE-PR): evidence of reliability and validity for caregivers reporting on children and adolescents with mental health concerns in Greece

**DOI:** 10.1101/2024.08.06.24311524

**Authors:** Konstantinos Kotsis, Graham Thornicroft, Julia Luiza Schafer, Aspasia Serdari, Maria Basta, Caio Borba Casella, Lauro Estivalete Marchionatti, Mauricio Scopel Hoffmann, Alexandra Tzotzi, Andromachi Mitropoulou, André Rafael Simioni, Katerina Papanikolaou, Anastasia Koumoula, Giovanni Abrahão Salum

## Abstract

**Background:** To improve access to mental health care for children and adolescents, it is necessary to identify the barriers faced by their caregivers. The aim of this study is to identify these barriers in Greece and to investigate the reliability and validity of the modified version of the Barriers to Access to Care Evaluation scale (BACE) - the BACE Proxy Report (BACE-PR).

**Methods:** A total of 265 caregivers completed the BACE-PR. Exploratory Factor Analysis (EFA) and Confirmatory Factor Analysis (CFA) were used to investigate the factor structure of the instrument. Item parameters were assessed via Item Response Theory. Interpretability was assessed by linking summed scores to IRT-based scores.

**Results:** Caregivers reported care costs as the major barrier to access. Obsessive compulsive symptoms and self-harm were the conditions for which caregivers reported the highest level of barriers. EFA and CFA suggested that a one-factor solution fit the data well (RMSEA = 0.048, CFI = 0.991, TLI = 0.990). Internal consistency was found to be high (ω=0.96). Average z-scores provided five meaningful levels of caregivers’ perceived barriers compared to the national average.

**Conclusions:** Caregivers face a variety of barriers to access mental health care for their children and this could partly explain the treatment gap in the Greek mental health sector. Our study provides evidence for the reliability and validity of the BACE-PR scale, which can aid to identify caregiver-perceived barriers and to design interventions to improve access to mental health care.

## Introduction

Nearly half of mental disorders emerge before the age of 18 [1]. It is well established that mental disorders are leading causes of disability and, if left untreated, may have a serious impact in educational achievement and social functioning [2–4]., Studies from all over the world, also suggest that a great proportion of children with treatable mental health disorder do not receive the treatment they need to get better [5–8]. This treatment gap highlights the significant barriers to accessing care within each country’s mental health system.. To increase the access to mental health care services for children and adolescents, it is crucial to understand the factors that facilitate or inhibit their ability to receive care.

In child and adolescent mental health, caregivers play a major role in facilitating help-seeking and often serve as the primary point of contact with mental health services [9, 10]. For in-person help seeking services, research has been found that family is the dominant influence for adolescents [11]. Caregivers, as key facilitators of their children’s entry into mental health care, may encounter various barriers, including difficulties in recognizing symptoms, attitudes and stigma towards mental health that influence decisions to seek treatment, and lack of knowledge about the appropriate type of care to look for [10, 12–15].

The literature on barriers to access mental health care is limited in at least two important ways. First, quantitative studies typically using lists of barriers for caregivers to indicate their presence or absence or to rate them on a Likert scale, usually lack validation [16–21]. Second, although tools for measuring self-reported barriers in the adult population have validation studies, they are usually limited to specific contexts or populations [22, 23].

One of the tools that have the potential to overcome some of these challenges is the Barriers to Access to Care Evaluation (BACE v3) [24]. The instrument may be used to identify key barriers to care experienced by individuals who currently use or have recently used secondary mental health services, and it shows potential utility for use with general population samples [24]. Moreover, it assesses changes in barriers following intervention programs. The BACE has been developed as a self-report measure for adults experiencing mental health conditions, still leaving a gap for the barriers encountered on children and adolescents’ mental health care.

Given this gap, as well as the fact that barriers may vary across countries due to cultural differences as well as differences in mental health system structure and facilities, the aim of the current study is twofold. First, we aim to investigate the children/adolescents caregivers’ main perceived barriers to access to mental health care in Greece. Second, we aim to test the psychometric properties of the adapted version of BACE - PR scale as a proxy measure exploring its validity to be used by caregivers of children in need of mental health care.

## Methods

### Participants

We used data from a 2022/2023 cross-sectional survey from the Child and Adolescent Mental Health Initiative (CAMHI) on the current state and needs for child and adolescent mental health in Greece based on multiple viewpoints [25]. A nationwide sample of 1,756 caregivers participated in the online survey, answering questions related to service use and access, literacy and stigma, parenting practices, and mental health needs of their children/adolescents. Out of them, 265 caregivers answered affirmatively to the question “*Does this child/adolescent have any mental health difficulty (any psychological problem, any problem with his behavior or learning) that you are aware of?*”. Subsequently they completed the BACE-PR scale. Recruitment occurred through an online respondent panel provided by the research company IQVIA OneKey. This panel was developed based on census quotas, reaching participants online via social media and website campaigns, search engine optimization, panelists’ friends referrals, and affiliate networks [26]. To avoid self-selection, the online surveys were automatically routed to respondents based on a specific algorithm. Data was collected and preserved according to the General Data Protection Regulation (GDPR) National Policy [27]. Ethical approval was granted by the Research Ethics Committee of the Democritus University of Thrace [approval number: ΔΠΘ/ΕΗΔΕ/42772/307].

### Instrument

The Barriers to Access to Care Evaluation (BACE) scale was developed in the Health Services and Population Research Department of the Institute of Psychiatry, Psychology and Neuroscience, King’s College, England [24]. The BACE scale was originally developed as a 30-item self-report instrument conceived to evaluate barriers to access to mental health. Originally, authors suggest two subscales, the stigma subscale consisting of 12 items and the non-stigma consisting of the 18 remaining items. In the current study we used a modified version, adapted for assessing barriers to care for children and adolescents as reported by caregivers. The King’s College London original lead author granted special consent for the adaptation.

#### BACE adaptation to be a proxy measure in Greece

BACE was culturally adapted and translated in Greek, following reported detailed guidelines of a five-stage cultural adaptation process (https://osf.io/crz6h/). Five stages included forward translation, synthesis of versions, back translation, expert committee review and pilot testing with population. The questions were modified to focus on children; eg “*Being unsure where to go to get professional care*” was modified to “*Being unsure where to go to get professional care for my child/adolescent*”. Work-related questions were modified to reflect child/adolescent contexts e.g. “*Concern about what people at work might think, say or do*” was modified to “ *Concern about what people at the school of my child/adolescent might think, say, or do*”. The process is described in detail in Karagiorga et al. [28]. As for the self-report (patient adult) measure, respondents (caregivers) should indicate whether each item has ever stopped or delayed or discouraged him/her from getting or continuing with mental health professional care for the child/adolescent they are responsible for (modified instruction to reflect children and adolescents). Scoring includes checking one of four possible answers: not at all (0), a little (1), quite a lot (2), or a lot (3) with higher scores indicating a greater barrier. For each barrier, according to authors, three different scores may be given; (a) the mean of the response scores, (b) the percentage reporting they have experienced the barrier to any degree (i.e. the % circling 1, 2 or 3) and (c) the percentage experiencing the barrier as a major barrier (i.e. the % circling 3).

### Statistical analysis

For the description of the BACE items we used the mean score for each item and percentages as suggested by authors of the original scale. To explore which mental health difficulties seem to face more barriers, we plotted a heatmap of the mental health difficulties versus each barrier (item) of the BACE scale, representing the proportion of participants with a mental health difficulty that experience each of the BACE listed barriers. We included only difficulties that have been reported in more than 5 participants, therefore delusions were excluded (N=2).

Our psychometric assessment included several steps. First, since there are no studies that adapted the BACE scale to a proxy version, we performed an Exploratory Factor Analysis (EFA) to explore its underlying structure by using the Maximum Likelihood Estimator (ML) and Geomin Oblique Rotation. The selection of factors was based on the scree plot of eigenvalues and on the factor loadings. After selection of the best model, a Confirmatory Factor Analysis (CFA) was performed to explore the fit of the model to our sample, by using the Pairwise Maximum Likelihood Estimator (PML). Model fit was evaluated with the following fit indices: the Comparative Fit Index (CFI), the Tucker-Lewis Index (TLI), the Root Mean Square Error of Approximation (RMSEA), and the Standardized Root Mean-square Residual (SRMR). A good fit is indicated by the following values: SRMR < 0.6; RMSEA < 0.06; TLI and CFI > 0.95 [29]. Reliability analysis was performed using Cronbach alpha and by Omega (ω) coefficient for our model. Cronbach alpha assumes equal loadings (essential tau equivalence) and a value of 0.7 is considered acceptable [30]. Omega estimates the proportion of variance in the observed total score attributable to all “modeled” sources of common variance. A value of >0.8 is considered strong [31].

For interpretability-the degree to which one can assign qualitative meaning to an instrument’s quantitative scores or change in scores [32]-BACE has polytomous response options and therefore the graded response model (GRM) was used to estimate item parameters. We also conducted unidimensional item response theory assessments to determine where BACE provides information according to the latent trait. Moreover, we estimated the IRT factor scores of the latent variable to rank them into percentiles aiming to provide a meaningful scoring to stakeholders and researchers.

Analysis was performed using the software RStudio version 2023.12.1 [32, 33] and the packages *lavaan* [34]*, psych* [35]*, ltm* [36] *, and semTools* [37]. Database sheets and the code is openly available at our repository (https://osf.io/crz6h/).

## Results

### Participants

Sample characteristics are shown in Table 1. The majority of the respondents were female (59.6%), and were in a relationship (83.0%). Nearly all participants (98.1%) have finished the mandatory (9 years) education in Greece. The majority of caregivers reported that their child had learning difficulties (N=95), attention-deficit/hyperactivity disorder (N=86) and anxiety (N=77).

**Table 1.** Caregiver characteristic and reported child diagnosis.

|  | **Mean** | **SD** |
| --- | --- | --- |
| Caregivers' age | 41.78 | 7.87 |
| Child's age | 11.69 | 4.07 |
|  | **n** | **%** |
| Gender (Female) | 158 | 59.6 |
| Relationship status |  |  |
| Single | 1 | 3.40 |
| Relationship/Cohabitation/Married | 220 | 83.0 |
| Separated/Divorced/Widowed | 36 | 13.6 |
| Educational Level |  |  |
| Mandatory | 4 | 1.51 |
| Non Mandatory | 98 | 37.0 |
| Higher | 162 | 61.1 |
| Other | 1 | 0.37 |
| Income |  |  |
| Less than 1000€ monthly | 89 | 33.6 |
| Between 1001 to 2,000€ monthly | 107 | 40.4 |
| Above 2000€ monthly | 58 | 21.9 |
| I don't know/ Not applicable | 11 | 4.1 |
| Child reported mental health difficulties by caregivers |  |  |
| Autism | 25 | 9.4 |
| Intellectual difficulties | 13 | 4.91 |
| Learning difficulties | 95 | 35.8 |
| Attention-deficit Hyperactivity Disorder (ADHD) | 86 | 32.5 |
| Excessive worries, or fears (anxiety) | 77 | 29.1 |
| Sadness, loss of pleasure and/or irritability (depression) | 26 | 9.8 |
| Headstrong and oppositional behaviors (conduct difficulties) | 32 | 12.1 |
| Obsessions and/or compulsions (Obsessive Compulsive Disorder) | 13 | 4.9 |
| Eating and weight problems | 28 | 10.6 |
| Delusions and hallucinations | 2 | 0.7 |
| Alcohol and drugs | 6 | 2.2 |
| Sleep difficulties | 38 | 14.3 |
| Self-harm and/or suicidal ideation/attempts | 5 | 1.8 |
| Nighttime enuresis (Bed wetting) | 13 | 4.9 |
| Other | 14 | 5.2 |

\*Total percentages of mental health difficulties exceed 100% since the item was a tickbox guiding caregivers to mark all that apply.

### Barriers to access to care: descriptive assessment

Mean scores of each item of BACE are presented in Table 2, while percentages reported each item as a barrier to any degree and as a major barrier are presented in Figure 1. All scores lie between 0.61 and 1.49 indicating that caregivers face overall barriers to a small degree. The barrier with the highest mean score is “*Not being able to afford the financial costs involved*” which concurrently is also the item with the highest percentage reporting this as a barrier to any degree as well as reporting it as a major barrier. Other highly reported barriers include the fear that the child might be seen as weak, the willingness of the caregivers to solve the difficulties by themselves, the uncertainty of where they should ask for help and the future consequences relating to job applications.

**Figure 1:**
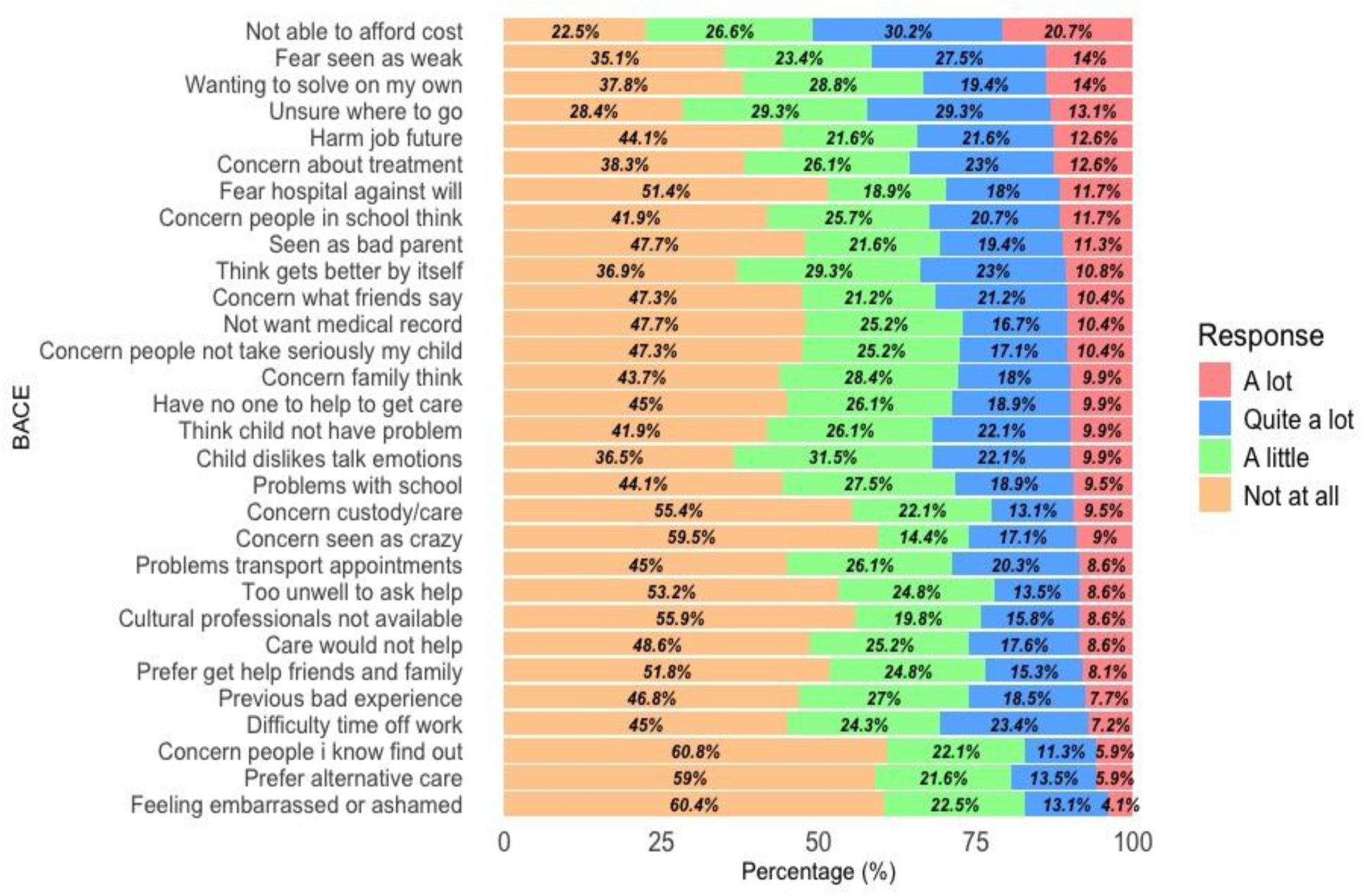
Caregivers ratings in each response (in percentage) of how much (“a lot”, “quite a lot”, “a little”, “not at all”) each item ever stopped, delayed or discouraged them from getting, or continuing with, professional care for a mental health problem. Descending order based on the percentage reported as a major barrier (circling “A lot”).

**Table 2.** Mean scores, frequencies and ranks for each barrier in the Barriers to Access to Care Evaluation Scale.

| No | Barrier | Mean | SD | % reporting barrier to any degree | % reporting as major barrier ('a lot') | Rank<br>(1 = item has highest proportion rating as a major barrier) |
| --- | --- | --- | --- | --- | --- | --- |
| 11 | Not being able to afford the financial costs involved. | 1.49 | 1.06 | 77.48 | 20.72 | 1 |
| 2 | Wanting to solve the problem of my child/adolescent on my own. | 1.09 | 1.06 | 62.16 | 13.96 | 2 |
| 3 | Concern that my child/adolescent might be seen as weak for having a mental health problem. | 1.2 | 1.07 | 64.86 | 13.96 | 3 |
| 1 | Being unsure where to go to get professional care for my child/adolescent | 1.27 | 1.02 | 71.62 | 13.06 | 4 |
| 5 | Concern that it might harm the child/adolescent's chances when applying for jobs in the future | 1.03 | 1.08 | 55.86 | 12.61 | 5 |
| 20 | Concerns about treatments available for my child/adolescent (e.g., medication side effects). | 1.1 | 1.05 | 61.71 | 12.61 | 5 |
| 4 | Fear of my child/adolescent being put in hospital against his/her will. | 0.9 | 1.08 | 48.65 | 11.71 | 7 |
| 28 | Concern about what people at the school of my child/adolescent might think, say, or do. | 1.02 | 1.05 | 58.11 | 11.71 | 8 |
| 14 | Concern that I might be seen as a bad parent. | 0.94 | 1.06 | 52.25 | 11.26 | 9 |
| 7 | Thinking the problem of my child/adolescent would get better by itself. | 1.08 | 1.02 | 63.06 | 10.81 | 10 |
| 19 | Concern that people might not take my child/adolescent seriously if they found out he/she was having professional care. | 0.91 | 1.03 | 52.7 | 10.36 | 11 |
| 21 | Not wanting a mental health problem to be on the medical records of my child/adolescent. | 0.9 | 1.03 | 52.25 | 10.36 | 12 |
| 26 | Concern about what the friends of my child/adolescent might think, say, or do. | 0.95 | 1.05 | 52.7 | 10.36 | 13 |
| 8 | Concern about what my child/adolescent's family might think, say, do or feel. | 0.94 | 1.01 | 56.31 | 9.91 | 14 |
| 18 | My child/adolescent dislikes talking about feelings, emotions, or thoughts. | 1.05 | 0.99 | 63.51 | 9.91 | 15 |
| 25 | Thinking my child/adolescent did not have a problem. | 1 | 1.02 | 58.11 | 9.91 | 16 |
| 30 | Having no one who could help me get professional care for my child/adolescent | 0.94 | 1.02 | 54.95 | 9.91 | 17 |
| 24 | Concern that the adolescent's children may be taken into care or that he/she may lose access or custody without his/her agreement. | 0.77 | 1.01 | 44.59 | 9.46 | 18 |
| 29 | Having problems with school while my child/adolescent receives professional care. | 0.94 | 1 | 55.86 | 9.46 | 19 |
| 12 | Concern that my child/adolescent might be seen as 'crazy'. | 0.76 | 1.04 | 40.54 | 9.01 | 20 |
| 6 | Problems with transport or traveling appointments. | 0.92 | 1 | 54.95 | 8.56 | 21 |
| 13 | Thinking that professional care for my child/adolescent would not help. | 0.86 | 0.99 | 51.35 | 8.56 | 22 |
| 15 | Professionals from the child/adolescent's own ethnic or cultural group not being available. | 0.77 | 1.01 | 44.14 | 8.56 | 23 |
| 16 | Being too unwell to ask for help for my child/adolescent. | 0.77 | 0.98 | 46.85 | 8.56 | 24 |
| 23 | Preferring to get help from family or friends for my child/adolescent. | 0.8 | 0.98 | 48.2 | 8.11 | 25 |
| 22 | Having had previous bad experiences with professional care for the mental health of my child/adolescent. | 0.87 | 0.97 | 53.15 | 7.66 | 26 |
| 27 | Difficulty taking time off work to take my child/adolescent to a health care service or professional. | 0.93 | 0.99 | 54.95 | 7.21 | 27 |
| 10 | Preferring to get alternative forms of care for my child/adolescent (e.g., traditional/religious healing or alternative/complementary therapies). | 0.66 | 0.92 | 40.99 | 5.86 | 28 |
| 17 | Concern that people I and my child/adolescent know might find out. | 0.62 | 0.9 | 39.19 | 5.86 | 29 |
| 9 | Feeling embarrassed or ashamed by my child/adolescent. | 0.61 | 0.86 | 39.64 | 4.05 | 30 |

Figure 2 presents the barrier heatmap according to each mental health difficulty reported. Obsessive compulsive disorder was the disorder that most caregivers faced the most barriers. Caregivers face almost every barrier for their children when they present with obsessions and compulsions. Other difficulties that face a lot of barriers in high degree are self-harm and intellectual disability. On the other hand, difficulties with a low level of barriers are mainly behavioral disorders as well as attention-deficit/hyperactivity disorder and autism spectrum disorder.

**Figure 2.**
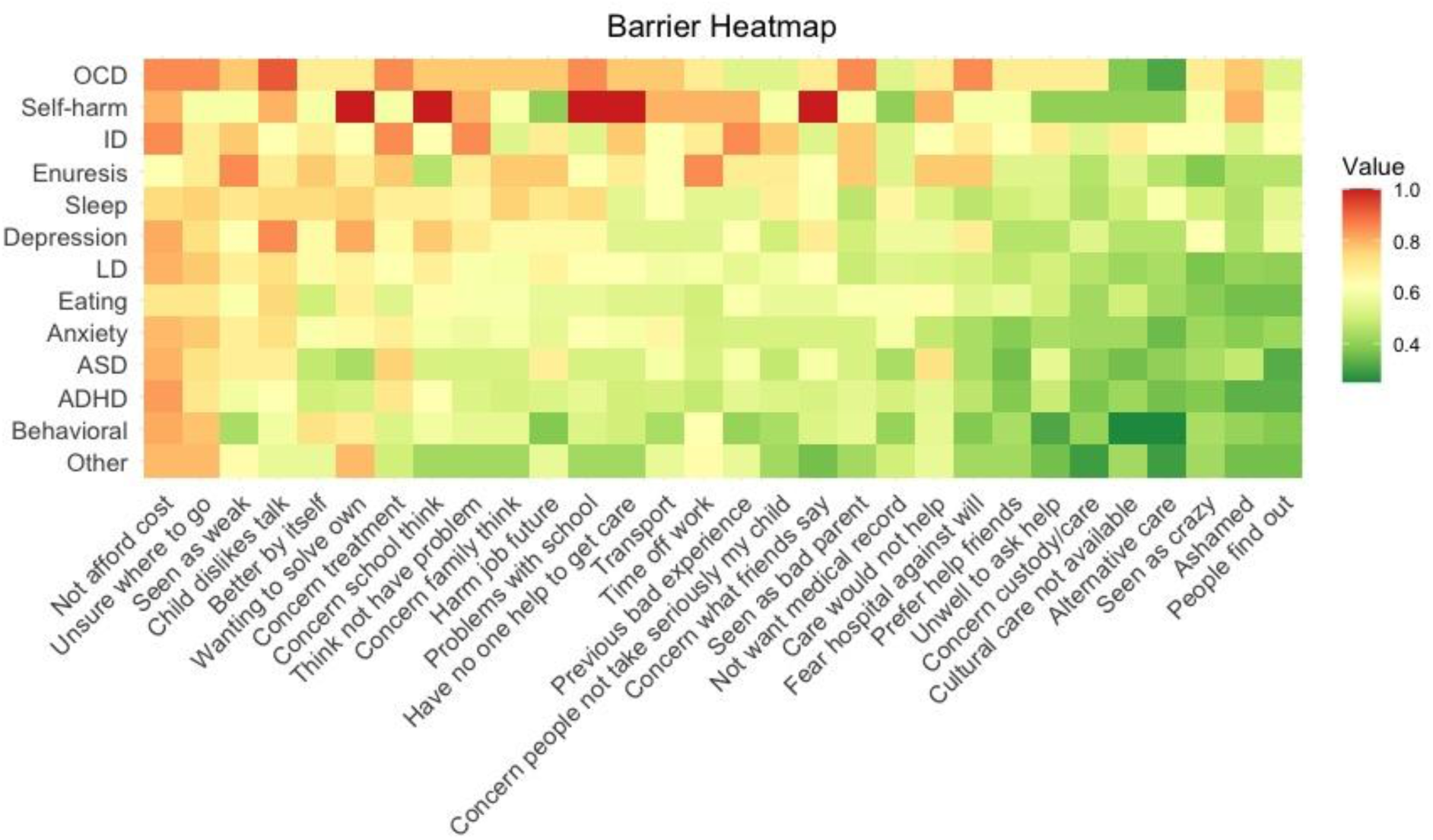
Percentage of caregivers reporting each barrier to any degree (answer: “A little”, “Quite a lot”, “A lot”) in each diagnosis. Order of BACE items according to percentage reported to any degree (descending left to right). Order of the mental health difficulties according to the overall level of barriers based on the mean BACE item scores (descending up to bottom). Mental health difficulties were plotted if N ≥ 5.

### Exploratory Factor Analysis

The results of the scree plot (<u>Figure S1</u>, Supplementary Material) favored a unidimensional solution. All 30 factor loadings were very high (>0.6) except two that displayed values of 0.38 and 0.59, although still significant.

### Confirmatory Factor Analysis

The unidimensional model (barriers to access to care) showed very good fit indices to the data (RMSEA = 0.048, CFI = 0.991, TLI = 0.990, SRMR = 0.061). Factor loadings were high, ranging from 0.38 to 0.89 (Table 3).

**Table 3.** Confirmatory Factor Analysis parameters and reliability coefficients.

| No | Item | Factor loadings | Threshold 1 | Threshold 2 | Threshold 3 |
| --- | --- | --- | --- | --- | --- |
| 1 | Being unsure where to go to get professional care for my child/adolescent | 0.509 | -0.594 | 0.179 | 1.127 |
| 2 | Wanting to solve the problem of my child/adolescent on my own. | 0.549 | -0.341 | 0.384 | 1.099 |
| 3 | Concern that my child/adolescent might be seen as weak for having a mental health problem. | 0.763 | -0.392 | 0.192 | 1.017 |
| 4 | Fear of my child/adolescent being put in hospital against his/her will. | 0.753 | 0.070 | 0.530 | 1.164 |
| 5 | Concern that it might harm the child/adolescent's chances when applying for jobs in the future | 0.786 | -0.146 | 0.392 | 1.109 |
| 6 | Problems with transport or traveling appointments. | 0.684 | -0.127 | 0.556 | 1.314 |
| 7 | Thinking the problem of my child/adolescent would get better by itself. | 0.660 | -0.314 | 0.393 | 1.150 |
| 8 | Concern about what my child/adolescent's family might think, say, do or feel. | 0.841 | -0.164 | 0.522 | 1.267 |
| 9 | Feeling embarrassed or ashamed by my child/adolescent. | 0.795 | 0.261 | 0.863 | 1.552 |
| 10 | Preferring to get alternative forms of care for my child/adolescent (e.g., traditional/religious healing or alternative/complementary therapies). | 0.835 | 0.209 | 0.842 | 1.429 |
| 11 | Not being able to afford the financial costs involved. | 0.375 | -0.728 | -0.029 | 0.850 |
| 12 | Concern that my child/adolescent might be seen as 'crazy'. | 0.835 | 0.256 | 0.631 | 1.257 |
| 13 | Thinking that professional care for my child/adolescent would not help. | 0.743 | -0.090 | 0.592 | 1.277 |
| 14 | Concern that I might be seen as a bad parent. | 0.757 | -0.024 | 0.550 | 1.273 |
| 15 | Professionals from the child/adolescent's own ethnic or cultural group not being available. | 0.720 | 0.127 | 0.654 | 1.333 |
| 16 | Being too unwell to ask for help for my child/adolescent | 0.799 | 0.078 | 0.709 | 1.352 |
| 17 | Concern that people I and my child/adolescent know might find out. | 0.894 | 0.256 | 0.867 | 1.404 |
| 18 | My child/adolescent dislikes talking about feelings, emotions, or thoughts. | 0.730 | -0.384 | 0.444 | 1.247 |
| 19 | Concern that people might not take my child/adolescent seriously if they found out he/she was having professional care. | 0.833 | -0.078 | 0.560 | 1.236 |
| 20 | Concerns about treatments available for my child/adolescent (e.g., medication side effects). | 0.684 | -0.324 | 0.342 | 1.140 |
| 21 | Not wanting a mental health problem to be on the medical records of my child/adolescent. | 0.743 | -0.084 | 0.572 | 1.266 |
| 22 | Having had previous bad experiences with professional care for the mental health of my child/adolescent. | 0.781 | -0.128 | 0.642 | 1.444 |
| 23 | Preferring to get help from family or friends for my child/adolescent. | 0.816 | 0.042 | 0.690 | 1.364 |
| 24 | Concern that the adolescent's children may be taken into care or that he/she may lose access or custody without his/her agreement. | 0.853 | 0.189 | 0.741 | 1.276 |
| 25 | Thinking my child/adolescent did not have a problem. | 0.733 | -0.234 | 0.430 | 1.239 |
| 26 | Concern about what the friends of my child/adolescent might think, say, or do. | 0.819 | -0.125 | 0.489 | 1.261 |
| 27 | Difficulty taking time off work to take my child/adolescent to a health care service or professional. | 0.654 | -0.114 | 0.448 | 1.346 |
| 28 | Concern about what people at the school of my child/adolescent might think, say, or do. | 0.771 | -0.217 | 0.439 | 1.201 |
| 29 | Having problems with school while my child/adolescent receives professional care. | 0.789 | -0.172 | 0.546 | 1.260 |
| 30 | Having no one who could help me get professional care for my child/adolescent | 0.793 | -0.110 | 0.568 | 1.350 |
| **Model Fit** |  |  |  |  |  |
|  | RMSEA | 0.048 |  |  |  |
|  | CFI | 0.991 |  |  |  |
|  | TLI | 0.990 |  |  |  |
|  | SRMR | 0.061 |  |  |  |
| **Reliability** |  |  |  |  |  |
| | McDonald's $\omega$ | 0.96 | | | |
|  | Cronbach's alpha | 0.96 |  |  |  |

Note: RMSEA= root-mean-square error of approximation; CFI=comparative fit index; TLI=Tucker–Lewis index; SRMR = standardized root-mean-square residual;

### Reliability

Unidimensional model presented with high reliability with values of 0.96 to McDonald’s ω as well as Cronbach’s alpha, indicating excellent internal consistency (Table 3).

### Interpretability

#### Unidimensional Item Performance Analysis

Item response function curves and item information curves for each item can be found in <u>Figures S3</u> and <u>S4</u>, supplemental material. Test information function plot (<u>Figure</u> S5) shows that BACE proxy version provides the most information about slightly-above-than-average barrier levels (the peak is around θ=1).

#### Linking summed score to IRT-based z-scores

Factor score from the IRT of the BACE proxy version as shown in Figure 3 follows the normal distribution. The z-scores for the latent variable (Table 4) provide a reference point to assess interpretability of the BACE. Based on those scores, we classified the amount of barriers that caregivers face as: (1) No barriers (BACE total = 0); (2) slight barriers (BACE total 1-31); (3) mild barriers (BACE total 32-44); (4) moderate barriers (BACE total 45-63); and (5) severe barriers (BACE total over 64).

**Figure 3.**
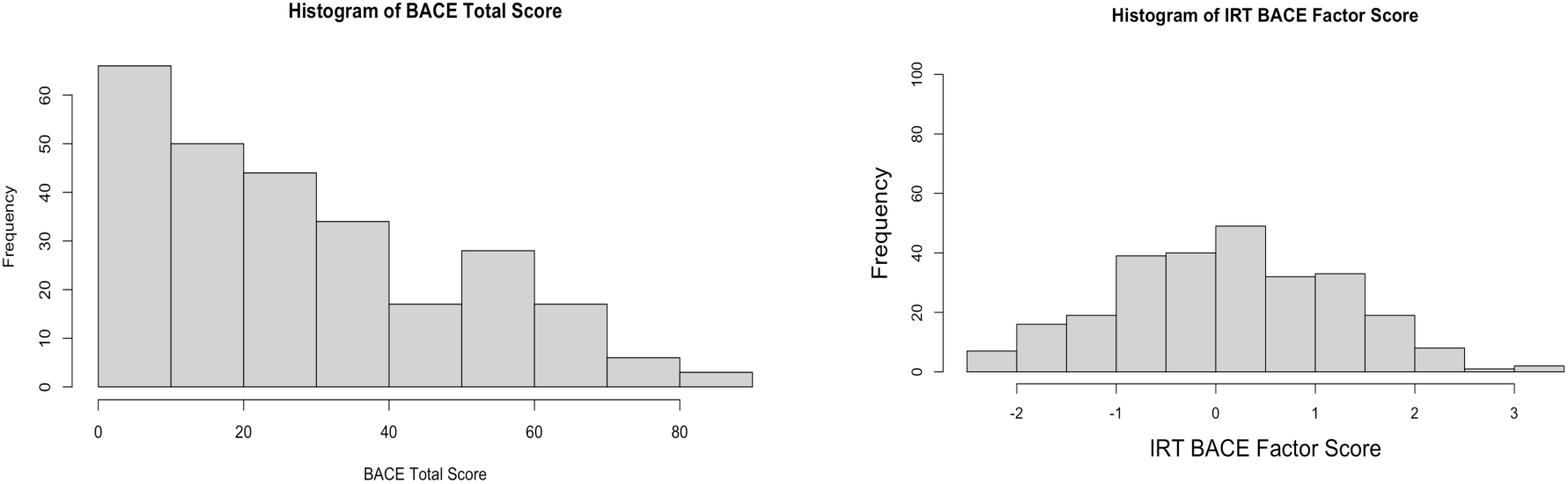
Histogram of the BACE Factor score from the Item Response Analysis

**Table 4.**
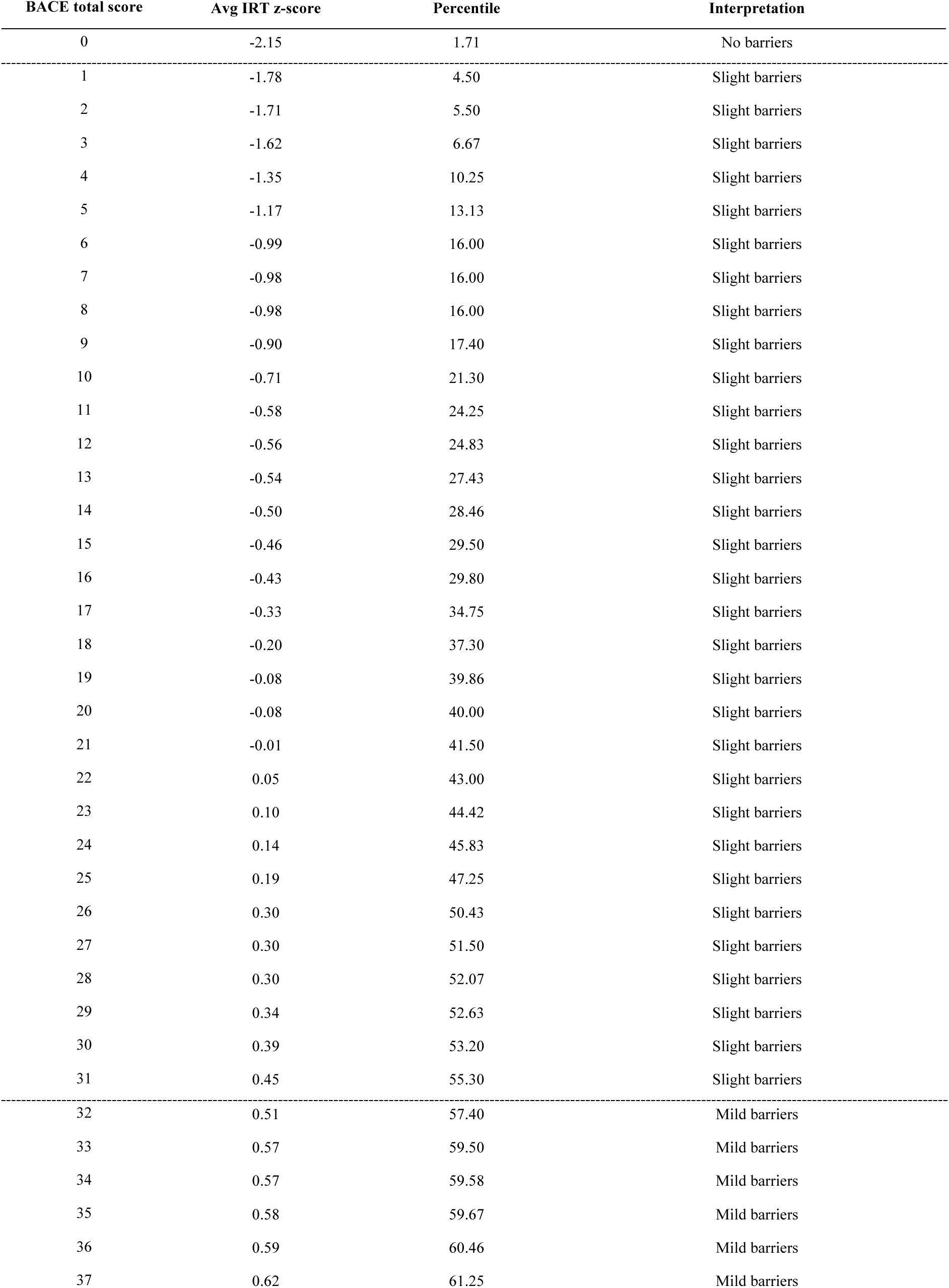
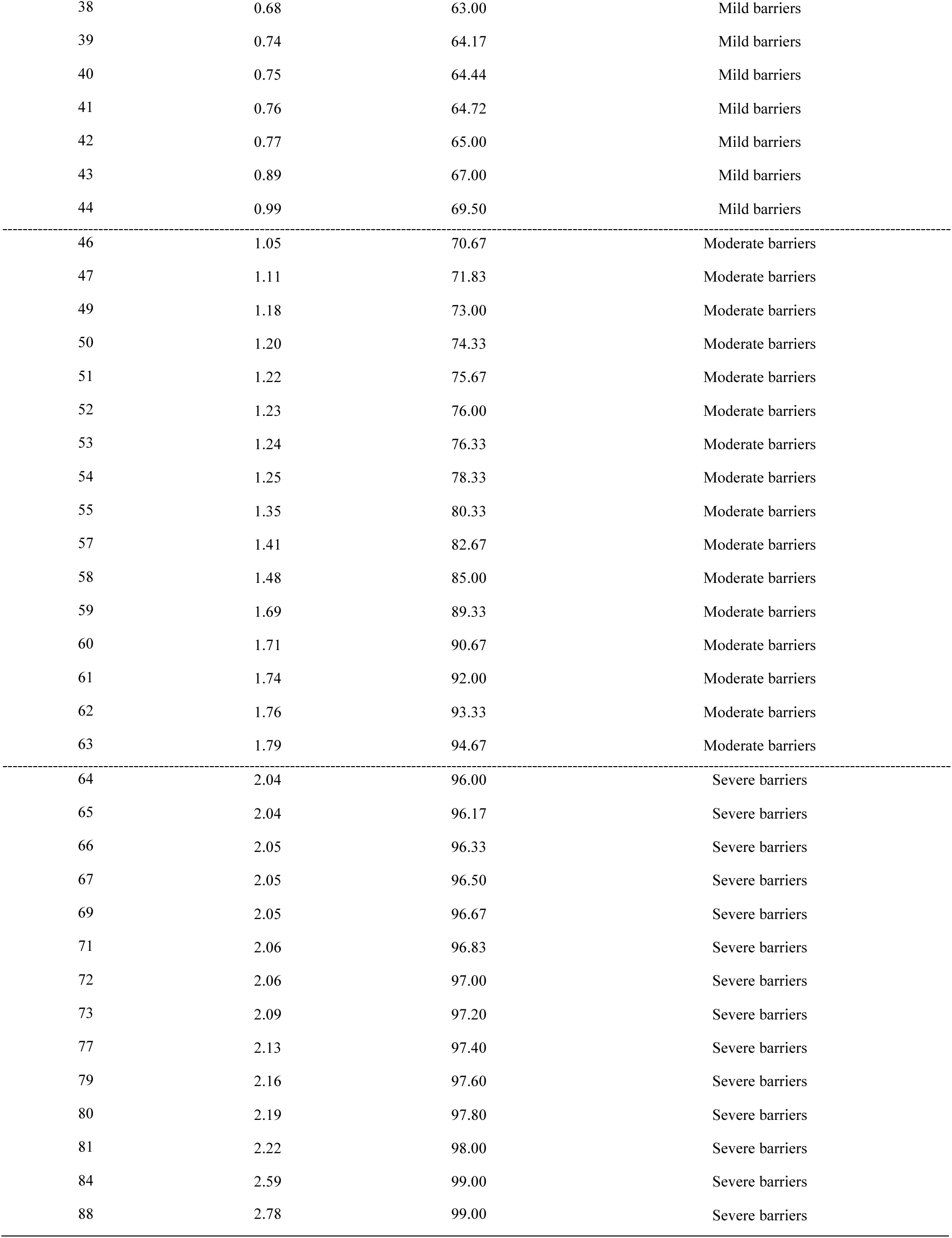
Interpretation of the Barriers to Access to Care Evaluation Scale total score.

| BACE total score | Avg IRT z-score | Percentile | Interpretation |
| --- | --- | --- | --- |
| 0 | -2.15 | 1.71 | No barriers |
| 1 | -1.78 | 4.50 | Slight barriers |
| 2 | -1.71 | 5.50 | Slight barriers |
| 3 | -1.62 | 6.67 | Slight barriers |
| 4 | -1.35 | 10.25 | Slight barriers |
| 5 | -1.17 | 13.13 | Slight barriers |
| 6 | -0.99 | 16.00 | Slight barriers |
| 7 | -0.98 | 16.00 | Slight barriers |
| 8 | -0.98 | 16.00 | Slight barriers |
| 9 | -0.90 | 17.40 | Slight barriers |
| 10 | -0.71 | 21.30 | Slight barriers |
| 11 | -0.58 | 24.25 | Slight barriers |
| 12 | -0.56 | 24.83 | Slight barriers |
| 13 | -0.54 | 27.43 | Slight barriers |
| 14 | -0.50 | 28.46 | Slight barriers |
| 15 | -0.46 | 29.50 | Slight barriers |
| 16 | -0.43 | 29.80 | Slight barriers |
| 17 | -0.33 | 34.75 | Slight barriers |
| 18 | -0.20 | 37.30 | Slight barriers |
| 19 | -0.08 | 39.86 | Slight barriers |
| 20 | -0.08 | 40.00 | Slight barriers |
| 21 | -0.01 | 41.50 | Slight barriers |
| 22 | 0.05 | 43.00 | Slight barriers |
| 23 | 0.10 | 44.42 | Slight barriers |
| 24 | 0.14 | 45.83 | Slight barriers |
| 25 | 0.19 | 47.25 | Slight barriers |
| 26 | 0.30 | 50.43 | Slight barriers |
| 27 | 0.30 | 51.50 | Slight barriers |
| 28 | 0.30 | 52.07 | Slight barriers |
| 29 | 0.34 | 52.63 | Slight barriers |
| 30 | 0.39 | 53.20 | Slight barriers |
| 31 | 0.45 | 55.30 | Slight barriers |
| 32 | 0.51 | 57.40 | Mild barriers |
| 33 | 0.57 | 59.50 | Mild barriers |
| 34 | 0.57 | 59.58 | Mild barriers |
| 35 | 0.58 | 59.67 | Mild barriers |
| 36 | 0.59 | 60.46 | Mild barriers |
| 37 | 0.62 | 61.25 | Mild barriers |
| 38 | 0.68 | 63.00 | Mild barriers |
| 39 | 0.74 | 64.17 | Mild barriers |
| 40 | 0.75 | 64.44 | Mild barriers |
| 41 | 0.76 | 64.72 | Mild barriers |
| 42 | 0.77 | 65.00 | Mild barriers |
| 43 | 0.89 | 67.00 | Mild barriers |
| 44 | 0.99 | 69.50 | Mild barriers |
| 46 | 1.05 | 70.67 | Moderate barriers |
| 47 | 1.11 | 71.83 | Moderate barriers |
| 49 | 1.18 | 73.00 | Moderate barriers |
| 50 | 1.20 | 74.33 | Moderate barriers |
| 51 | 1.22 | 75.67 | Moderate barriers |
| 52 | 1.23 | 76.00 | Moderate barriers |
| 53 | 1.24 | 76.33 | Moderate barriers |
| 54 | 1.25 | 78.33 | Moderate barriers |
| 55 | 1.35 | 80.33 | Moderate barriers |
| 57 | 1.41 | 82.67 | Moderate barriers |
| 58 | 1.48 | 85.00 | Moderate barriers |
| 59 | 1.69 | 89.33 | Moderate barriers |
| 60 | 1.71 | 90.67 | Moderate barriers |
| 61 | 1.74 | 92.00 | Moderate barriers |
| 62 | 1.76 | 93.33 | Moderate barriers |
| 63 | 1.79 | 94.67 | Moderate barriers |
| 64 | 2.04 | 96.00 | Severe barriers |
| 65 | 2.04 | 96.17 | Severe barriers |
| 66 | 2.05 | 96.33 | Severe barriers |
| 67 | 2.05 | 96.50 | Severe barriers |
| 69 | 2.05 | 96.67 | Severe barriers |
| 71 | 2.06 | 96.83 | Severe barriers |
| 72 | 2.06 | 97.00 | Severe barriers |
| 73 | 2.09 | 97.20 | Severe barriers |
| 77 | 2.13 | 97.40 | Severe barriers |
| 79 | 2.16 | 97.60 | Severe barriers |
| 80 | 2.19 | 97.80 | Severe barriers |
| 81 | 2.22 | 98.00 | Severe barriers |
| 84 | 2.59 | 99.00 | Severe barriers |
| 88 | 2.78 | 99.00 | Severe barriers |

## Discussion

The aim of this study was to identify the barriers that caregivers face to access mental health care in Greece for their children and to explore the reliability and validity of the culturally adapted BACE-PR in a nationwide sample of caregivers with mental health concerns of their children. The top five major barriers to access care in Greece were "Not being able to afford the financial costs involved", "Wanting to solve the problem of my child/adolescent on my own’’, "Concern that my child/adolescent might be seen as weak for having a mental health problem", "Being unsure where to go to get professional care for my child/adolescent" and "Concern that it might harm the child/adolescent’s chances when applying for jobs in the future". We showed consistent evidence for the reliability and validity of BACE-PR as a unidimensional construct. We also provide practical recommendations for interpretability of the BACE-PR by means of using IRT based scores (z-scores).

One of the most important findings of this study is that three quarters of the caregivers reported financial costs as a barrier for accessing mental health care. This is likely to relate to the consequences of 12 years of continuous financial crisis in Greece, which seemed to be worsened by the increasing demands of the sector after the COVID-19 pandemic. During the crisis years, unemployment rates were increased and loss of income was noted in the majority of professions. Moreover, mental health is known to be particularly vulnerable to rapid economic fluctuations [38–40]. Therefore, an increase in demand for mental health services was noted, while at the same time public mental health services operated with fewer employees with long waiting lists [41]. Although the public mental health system is mainly free of charge, many caregivers look for care in the private sector. However, the cost of private treatment for children with mental and neurodevelopmental disorders is only partially covered by the national insurance fund, thus caregivers often have to pay out-of-pocket costs for their child’s treatment. Our finding is consistent with the international literature where in many studies the cost represents a major barrier [42–44].

Another significant finding of our study is that a set of barriers to mental health access among caregivers of children with mental health problems in Greece are related to attitudes and stigma. Several studies in the literature suggest that these types of barriers are noted among caregivers [45–47]. To our knowledge, up to now, no relevant studies exploring caregivers’ beliefs on their children’s mental health services barriers have been conducted in Greece. However, stigma for mental illness is prevalent in Greek culture according to a systematic review [48]. Finally, the lack of knowledge about where to refer for help also represents a barrier in our study, consistent with the literature [47, 49, 50]. This may reflect the fragmentation of the Greek mental health system and the overall challenges the system faces in Greece [51, 52]. Current initiatives are trying to mitigate those barriers by making information easily accessible to the Greek population (www.camhi.gr).

Based on our study, in Greece caregivers of children with obsessive compulsive symptoms (OCS), self-harm behaviors, or intellectual disability (ID) seem to face the most barriers and to a higher level compared to caregivers of children with other disorders in our sample. These conditions are severe, often associated with severe impairment in everyday functioning and may need specialized services. Parents of children with OCD found themselves caught in “loops” in which they engaged in repeated process steps due to the emergence of barriers, when they assess exposure therapy. They also feel isolated and guilty because of the burden of finding treatment for their children [53]. Moreover, poor mental health literacy and stigmatizing attitudes are considered main parameters for the delayed help seeking behavior in OCD [54]. Similar reasons seem to apply to help seeking for young people at risk of self-harm [55, 56]). It is well known that stigma is high for youth’s self-harm behaviors and the care they receive in the emergency departments is often inadequate [56–58]. Caregivers of children with ID also indicate they face high levels of barriers that might be related to the barriers they also face in educational settings. Furthermore, when ID is comorbid with other disorders, such as autism spectrum disorders (ASD), it increases the likelihood of unmet mental health care needs [59]. Moreover, in the case of Greece we should also highlight structural barriers such as the lack of specialized services providing evidence-based treatment for these conditions, especially in the public sector. On the other hand, our findings showed that behavioral disorders and other neurodevelopmental disorders such as autism spectrum disorder and attention deficit hyperactivity disorder face a lower level of barriers when compared to other mental health concerns. We hypothesize that externalizing problems as well as communication deficits and social impairment in ASD are more likely to prompt caregivers to ask for help and that there are many private services in Greece that provide care for these conditions.

We also found that the psychometric testing of the BASE-PR provides good evidence for data quality and internal consistency. The unidimensional model fits well the data indicating that the BACE-PR can be used to measure the barriers that the caregivers face when they need to access mental health care for their children. The unidimensional solution seems in contrast with the literature considering that the original version of the BACE divides the items as stigma non-related and stigma [24], which was corroborated by the Arabic, the Japanese (excluding 6 items and with marginal fit indices) and the French versions [60–62]. Our unique finding of unidimensionality might be explained by the fact that our tool represents a modified proxy version. Based on that, we suggest that the BACE-PR scale should be treated as a measure of the overall perceived barriers. In terms of practicality and interpretability, the results of this study can be used by stakeholders to understand the quality and quantity of the barriers caregivers face. Reliability of our proxy version was also found very high, consistent with the above-mentioned translations.

### Limitations and strengths

Our study has important limitations. First, the sample size is small to conceptualize which barriers caregivers face in Greece. This also prevented us from doing EFA and CFA in distinct samples, as recommended by the literature for psychometric assessment of psychological instruments [63, 64]. Second, this is an online panel based sample, thus implying representative bias for groups without internet access. Our study also has strengths that should be highlighted. To the best of our knowledge this is the first study using a modified version of BACE, providing to the international community a tool to assess barriers to access care in children and adolescents. In addition, it is the first study in Greece exploring the barriers that caregivers face in access to mental health care. Second, we followed a rigorous cross-adaptation process for the instrument. Third, item performance analyses in a nationwide survey, provided psychometric evidence of BACE-PR adequacy on an item-based approach, surpassing the limitations of classical test theory analyses. Finally, our study provides to the Greek service providers and stakeholders a valid tool to record caregivers’ perceived barriers to design interventions to address them.

### Conclusions

In this study, we developed and validated the BACE-PR tool, addressing a gap of tools for caregiver report of barriers to child and adolescent mental health care. Caregivers of children with mental health concerns may encounter numerous barriers when help from mental health professionals is needed. Identifying them is important to understand the treatment gap at both the national and at the local level considering the different needs across the country, especially in terms of the uneven distribution of services and professionals. The present study highlights the need for free of charge specialized interventions to address the financial barriers. Moreover, interventions to fight stigma and to guide caregivers on how to access the available services (e.g. with detailed service mapping) are necessary in Greece. To improve access to treatment and to provide early and comprehensive services, we need also to explore variations in the perceived barriers among caregivers of children across different mental health disorders and different ages. Finally, the present study supports the use of BACE-PR to identify perceived barriers. Future studies in larger scale populations based on this tool are needed to confirm our findings. Furthermore, appropriate interventions may be applied by policy makers to address the barriers and longitudinal studies may evaluate their effect on childrens’ mental health in Greece. Moreover, specifically addressing barriers at a more regional level can also be a positive strategy for fighting barriers considering the different needs across the country, especially in terms of the uneven distribution of services and professionals. Diminishing the barriers and therefore increasing access to mental health care would be extremely beneficial for children and adolescents, since it will allow for early intervention and overall decrease the treatment gap.

## Data Availability

The dataset statistical codes supporting the conclusions of the current study are openly available in the CAMHI Open Science Framework repository [http://doi.org/10.17605/OSF.IO/CRZ6H]

http://doi.org/10.17605/OSF.IO/CRZ6H

## Acknowledgments

The Stavros Niarchos Foundation (SNF) under its Global Health Initiative has partnered with the Child Mind Institute and a countrywide Network of mental health providers in the public system to jointly design, launch, and deliver the Child and Adolescent Mental Health Initiative (CAMHI) in Greece, which supports this work. The authors would like to thank SNF’s Co-President Andreas C. Dracopoulos and CMI’s President Dr. Harold Koplewicz for their leadership in creating, launching, and supporting this project. We would also like to thank Ms. Elianna Konialis, Ms. Dimitra Moustaka and Mr. Panos Papoulias for their critical role in multiple steps of the conceptualization and implementation of the CAMHI objectives. Finally, thanks to CAMHI hub leaders and hub members that participated in multiple stages of this work and participants and their families for their contribution.

## Declarations

### Availability of data and materials

The dataset is openly available in the CAMHI Open Science Framework repository [http://doi.org/10.17605/OSF.IO/CRZ6H]

### Code availability

The statistical codes supporting the conclusions of the current study are openly available in the CAMHI Open Science Framework repository [http://doi.org/10.17605/OSF.IO/CRZ6H]

### Authors’ contribution

KK and GS have drafted the work. Data analysis was performed by KK and GS. Material preparation and data curation were performed by KK, JS, LM, AT, AM, GT and AS. Formal analysis was performed by KK, MH and GS. Project administration was performed by KK, KP, MB, AS, AK and GS. GS supervised the work. All authors contributed to data presentation; visualization by KK, CC and GS. All authors substantially contributed to interpretation of data, reviewed and edited the work. All authors read and approved the final manuscript. All authors have agreed both to be personally accountable for the author’s own contributions and ensure that questions related to the accuracy or integrity of any part of the work, even ones in which they are not personally involved, are appropriately investigated, resolved, and the resolution documented in the literature.

### Ethical approval

Ethical approval was granted by the Research Ethics Committee of the Democritus University of Thrace [approval number: ΔΠΘ/ΕΗΔΕ/42772/307].

### Consent to participate

Informed consent was obtained from all individual participants included in the study.

### Consent for publication

Not applicable

