## Supplemental Material for "Barriers to Access to Care Evaluation Scale - Proxy Report (BACE-PR): evidence of reliability and validity for caregivers reporting on children and adolescents with mental health concerns in Greece"

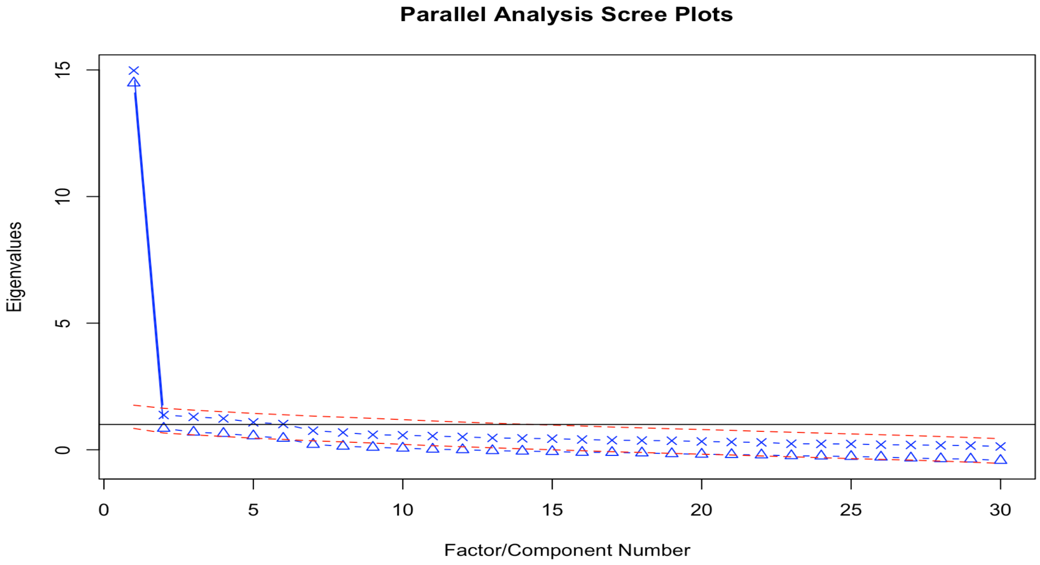


**Supplemental Figure S1:**Parallel analysis plot

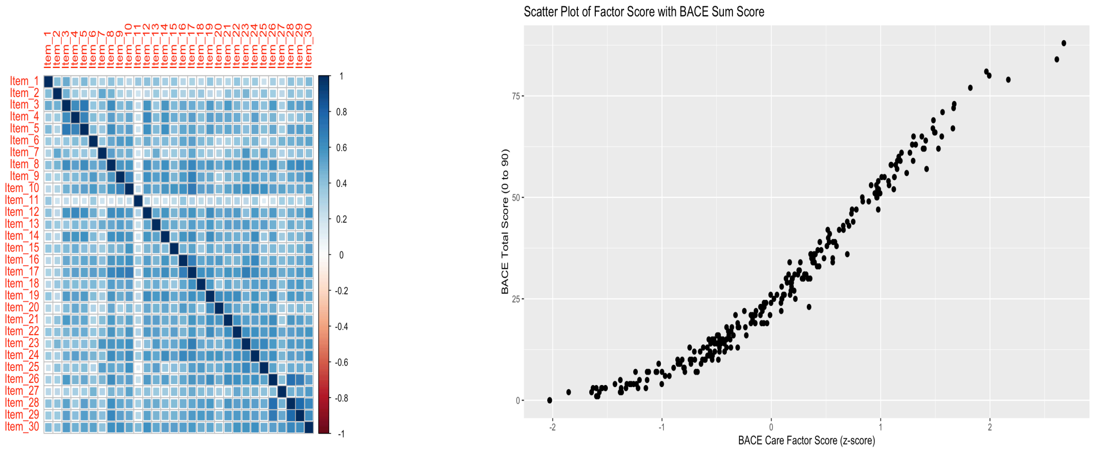


**Supplemental Figure S2.** Correlation matrix and scatter plot showing the association between summed score and IRT-based score

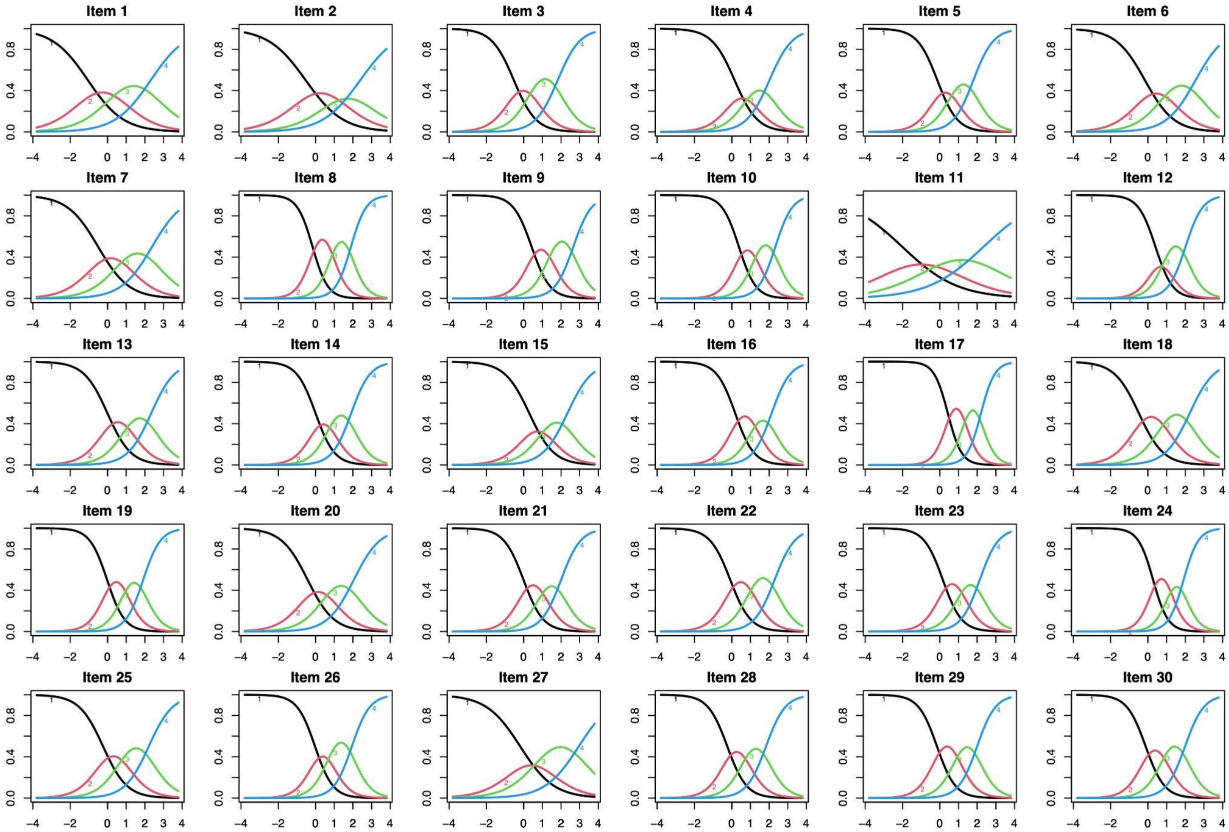


**Supplemental Figure S3:** Item Response Characteristic Curves (unidimensional solution)

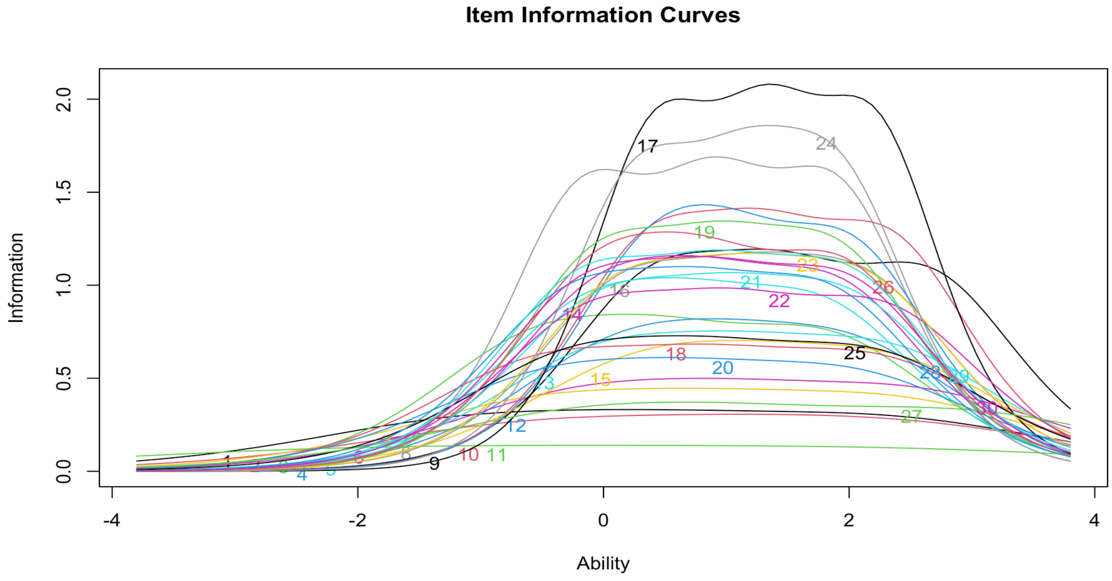


**Supplemental Figure S4.** Item Information Curves (unidimensional solution)

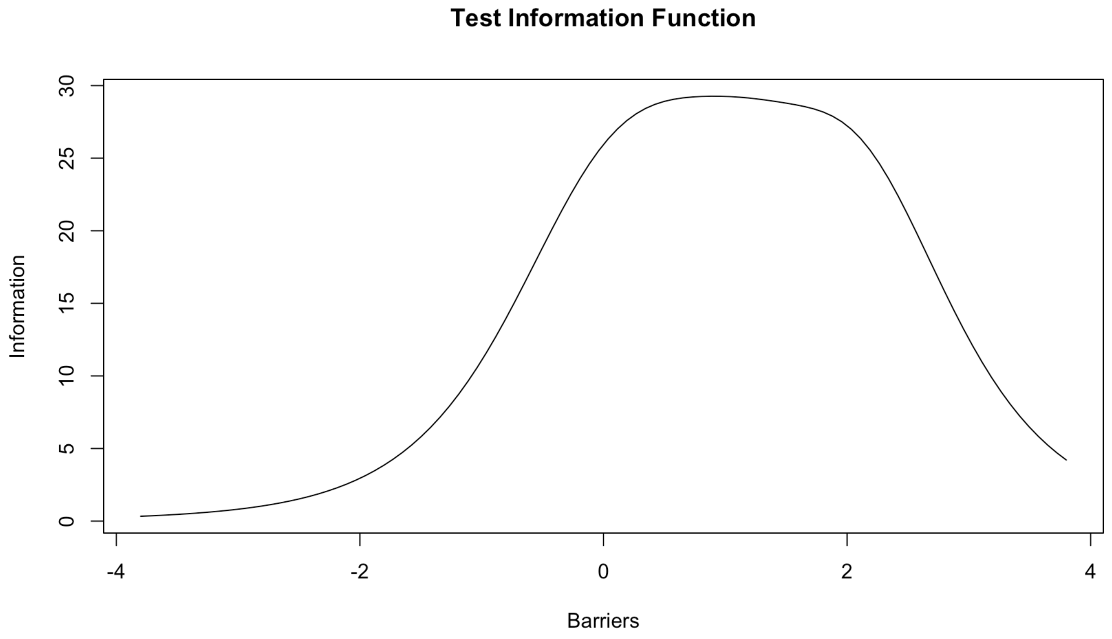


**Supplemental Figure S5.** Test Information Function of the Barriers to Access to Care Evaluation (unidimensional solution)
